# Symptoms and risk factors for hospitalization of COVID-19 presented in primary care

**DOI:** 10.1101/2021.03.26.21254331

**Authors:** S Rabady, K Hoffmann, M Brose, O Lammel, S Poggenburg, M Redlberger-Fritz, K Stiasny, M Wendler, L Weseslindtner, S Zehetmayer, G Kamenski

**Affiliations:** MD, Karl Landsteiner University of Health Sciences, Department of General Health Studies, Division General and Family Medicine, Dr. Karl-Dorrek-Straße 30, 3500, Krems, Austria; Assoc. Prof. Priv.-Doz. Dr., MPH, Associate Professor, Head of Unit Health Services Research and Telemedicine in Primary Care, Department of Preventive- and Social Medicine, Center for Public Health, Medical University of Vienna, Kinderspitalgasse 15, 1090 Vienna, Austria; MD, Praxis Dr Oliver Lammel, Ramsau am Dachstein; MD, Ordination Dr. Stephanie Poggenburg, Hart bei Graz; MD, Doz.Dr, Center of Virology, Medical University Vienna, Vienna, Austria; Prof, Center of Virology, Medical University Vienna, Vienna, Austria; Karl Landsteiner University of Health Sciences, Department of General Health Studies, Division General and Family Medicine, Dr. Karl-Dorrek-Straße 30, 3500, Krems, Austria; Assoc.Prof. Dr, Center for Medical Statistics, Informatics and Intelligent Systems, Medical University of Vienna, Spitalg.23 1090 Vienna; MD, Karl Landsteiner University of Health Sciences, Department of General Health Studies, Division General and Family Medicine, Dr. Karl-Dorrek-Straße 30, 3500, Krems, Austria; Karl Landsteiner Institute for Systematics in General Medicine, Angern, Austria

**Keywords:** Primary Care, COVID-19, symptoms, hospital admission

## Abstract

**Objective:** To extend knowledge of early symptoms as a precondition of early identification, and to gain understanding of associations between early symptoms and the development of a severe course of the disease.

**Design:** Retrospective observational study

**Setting:** Austrian GP practices in the year 2020, patients above 18 years were included.

**Participants:** We recruited 22 practices who included altogether 295 participants with a positive SARS-CoV-2 test.

**Main outcome measures:** Data collection comprised basic demographic data, risk factors and the recording of symptoms at several points in time in the course of the illness. Descriptive analyses for possible associations between demographics and symptoms were conducted by means of cross table. Group differences (hospitalized yes/no) were assessed using Fisher’s exact test. The significance level was set to 0.05; due to the observational character of the study, no adjustment for multiplicity was performed.

**Results:** Little more than one third of patients report symptoms generally understood to be typical for Covid-19. Most patients present with a variety of unspecific complaints. We found symptoms indicating complicated disease, depending on when they appear. The number of symptoms is likely to be a predictor for the need of hospital care. More than 50% of patients still experience symptoms 14 days after onset.

**Conclusions:** Underrating unspecific symptoms as possible indicators for SARS-CoV-2 infection harbours the danger of overlooking early disease. Monitoring patients during their illness using the indicators for severe disease we identified may help to identify patients who are likely to profit from early intervention.

**Data availability statement:** All data referred to in the manuscript are available from: Department of General Medicine and Family Practice, Karl Landsteiner Privatuniversitaet, Krems, Austria

**Article Summary:** *Strengths and limitations:*

- This study investigates data on the course of COVID-19 collected exclusively from patients in primary care and explores a wide range of symptoms.
- GPs were free to make their own testing decision according to their clinical judgement, and they followed each patient individually from day 1 to day 10 or 14.
- Limitations of our study concern the limited number of patients, due to the increased workload under difficult working conditions during the pandemic as well as the effort not being remunerated. However, the number of cases needed to identify group differences was calculated in advance, and this number has been reached. Our overall results are in accordance with our preliminary result analyses.

## Introduction

A central aspect in the containment of the COVID-19 pandemic is identification and isolation of possibly infectious persons, to prevent further spreading of the disease. Several studies were conducted with the goal of identifying diagnostic criteria that allow clinical differentiation between Covid- and non-Covid-infections: Most investigations used data collected from hospitalized patients (1–6); thus, from patients with severe disease. These studies have found high prevalence of fever (around 90%), dyspnoea (up to 50%), cough (60-70%), and fatigue in patients with COVID-19. Several other studies evaluated self-reported data from symptom tracker apps or outpatient clinics (2, 7–10). We could find but one investigation including additional data derived from primary care health records (9). Studies conducted in non-hospitalized patients reported a lower prevalence of the symptoms mentioned above, and a wide spectrum of additional symptoms, like myalgia, rhinorrhoea and/or nasal congestion, headache, sore throat, gastrointestinal and cardiovascular disturbances (7–9, 11). Loss of taste and/or smell was found to be rather specific when present (8, 11). All patients included in those studies had gone through a selection process before testing – by case definitions and testing criteria, by epidemiological factors or by previous tests (e.g. imaging). Some of these studies included only patients that had tested positive (10), others investigated patients that had tested positive or negative (8, 9). Patients not fulfilling established criteria may have escaped testing, and the symptoms found may reflect testing criteria (7). Some authors suspect that possibly a large proportion of COVID-19 cases are never tested and, thus, never recorded (12). This has not been investigated so far.

### What this study can add

Patients presenting with other than the canonical symptoms might be overlooked by current testing strategies and screening tools. Patients with discreet and seemingly unsuspicious complaints tend to be mobile and can widely spread the disease. Awareness among stake holders as well as in the general public as to the wide range of uncharacteristic symptoms is most needed to promote low-threshold and high sensitivity testing, and to advise repeated testing if any symptoms are present. This seems a requirement for effective containment strategies.

Austrian GPs are entitled to make an individual testing decision according to clinical judgement like when there is no alternative explanation for the symptom presented. Austrian GP practices can send their own samples for SARS-CoV-2 PCR to be analysed via a Surveillance Network based at the Medical University Vienna, and from mid-October 2020 point of care testing for SARS-CoV-2 in GP`s offices has become possible. 25% of our study practices were part of this network. GP practices in Austria are easy to access; in general, it is possible to walk in or to get an appointment on the same day.

Against this background, it was the aim of this study to assess early COVID-19 symptoms and their development in patients of different demographic groups in primary care as well as their possible associations with complications in the course of the disease.

## Patients, Materials and Methods

This study was designed as an observational, retrospective study in General Practice in Austria. Recruitment of practices and participants took place between July 2020 and December 2020.

The Austrian Society of General Practice invited their members, publicly funded GPs and their practices, to participate. For this purpose, first announcements and invitation letters were sent out between April and July 2020. After receiving a positive vote by the Ethics Committee of the Karl-Landsteiner University for Health Sciences, practices interested in participating were informed about the aims of this study in detail and after agreement study material was provided.

Participating GPs included patients above 18 years either after testing positive at the point of care or after reporting to their GP with a positive PCR test from another testing facility. These persons were invited to participate in this study. If they were willing to participate and after the provision of their written informed consent, they were included in the study.

### Study material and data collection

A questionnaire using the open-source CDC program Epi Info 7 was designed to record demographic and anamnestic data, comorbidities, medication groups and risk factors regarding COVID-19 (Annex). Data extracted from the electronic health records (EHR) of the practices were transferred to the questionnaire. In addition, clinical parameters for assessing the patients` health status over a period of two weeks starting with the date of the positive test result as day 1, were documented. Additional assessment days were day 5, 7, 8, 10 and 14. Clinical data were self-reported in a monitoring sheet either by the patients themselves, or acquired via phone calls by the GPs. Data regarding patients` health status and symptoms were temperature, blood pressure, heart rate, dyspnoea, chest pressure, tightness of chest, malaise, weakness, headache, rhinitis, anosmia, ageusia, sore throat or gastrointestinal symptoms (Annex 2). These clinical parameters were selected using published studies of signs and symptoms of patients with COVID-19 (13, 14). All data were pseudonymized before forwarding them to the study centre. There, the data were checked for completeness and correct entries. Ambiguities were clarified by telephone call or e-mail contact with the participating GP practice.

### Data analysis

Data were converted into Excel files and accompanying statistical analysis was done using the statistical software program R (version 3.5.1).

Participants’ demographics as well as symptoms were first analysed descriptively. Descriptive analyses for possible associations between demographics and symptoms were conducted by means of cross tables. Group differences (hospitalized yes/no) were assessed using Fisher’s exact test. The significance level was set to 0.05; due to the observational character of the study, no adjustment for multiplicity was performed.

## Results

Altogether, 25 GP practices and 295 patients from seven Austrian federal states could be recruited. On average, the practices recruited 12 patients (SD 8.94, min 2 - max 31).

As shown in table 1 in detail, slightly more women than men were included in the study. In addition, the percentage of obese persons (BMI >30) was 19,0 %, which is slightly higher than the general Austrian average of 16% (15). On the other hand, only 7% of participants were smokers which is less than half of the Austrian average of 20%. (16).

### Initial symptoms, and development of symptomatology

The most common out of the 13 symptoms to be selected were joint or muscle pain and malaise on day 1, each of them reported by half of the patients (Table 2, Fig.1). Loss of smell/taste was reported on day 1 by less than a quarter of patients but became the most frequently expressed complaint from day 7 onwards until the end of the observational period on day 14. Fatigue was the 3rd most prevalent symptom on day 1, and the most common symptom on day five, and was still highly prevalent on day 10. So, fatigue was found to be the most persistent of symptoms of all. Cough turned out to be a less common symptom on day 1 (5th of 13 symptoms), becoming more frequent from day 5 onwards. Fever was reported by one third of participants as an initial symptom.

### Associations between symptoms and hospitalization

Analysis for associations of symptoms with the likelihood of hospitalization yielded the following results: For presence of fever and/or malaise from day 5 onwards we found significant associations with the need for hospitalization sometime in the further course of the disease. Headache showed a significant association if present on day 10. The likelihood for hospitalization was significantly increased in patients with either dyspnoea, fatigue, tightness of chest, and cough. In contrast, persons with rhinitis, sore throat, chest pain and anosmia as an initial symptom were less likely to need hospital care (Table 3, Fig.2).

**Figure 2.**
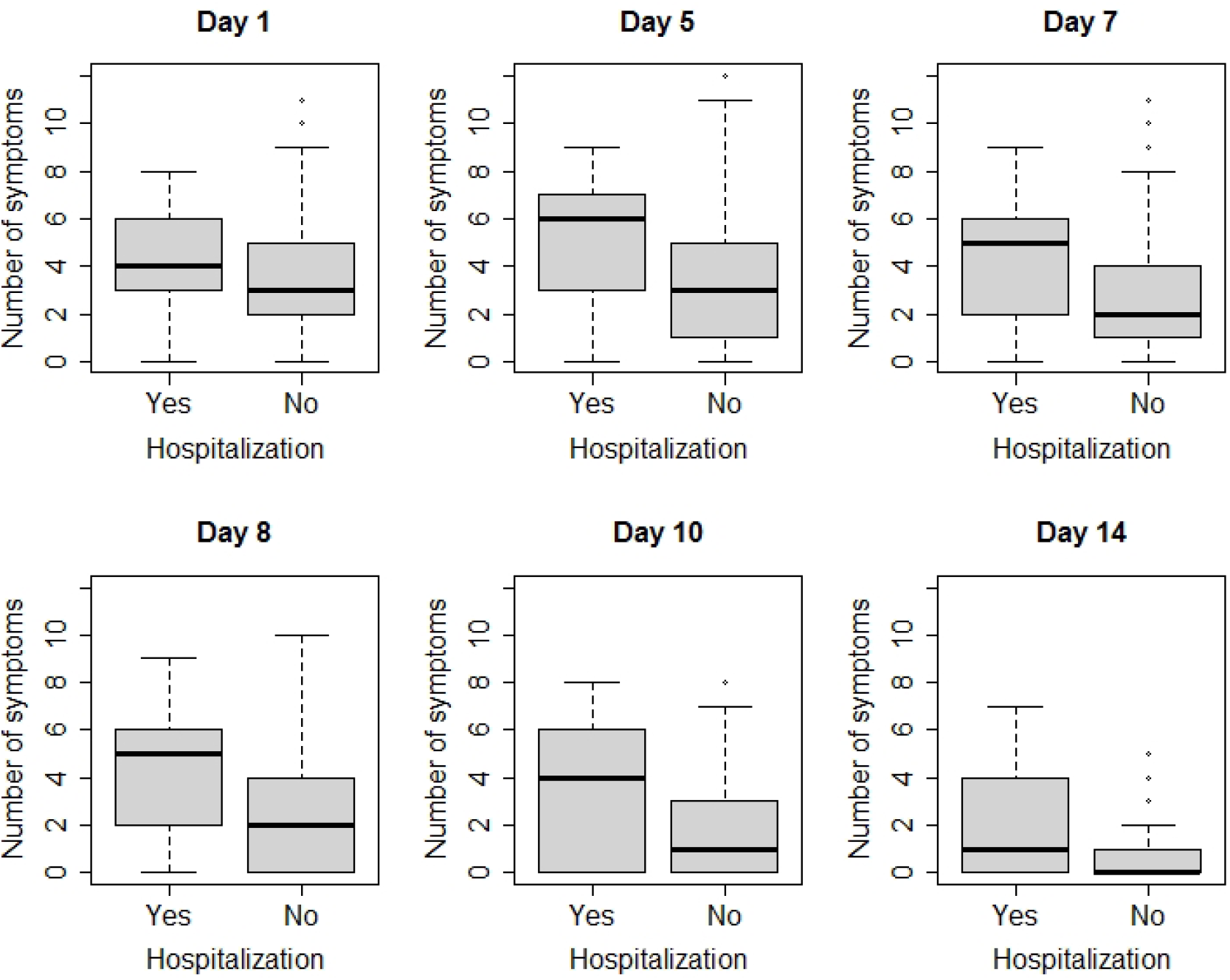
Number of symptoms in comparison of hospitalized to non-hospitalized patients

**Table 3:**
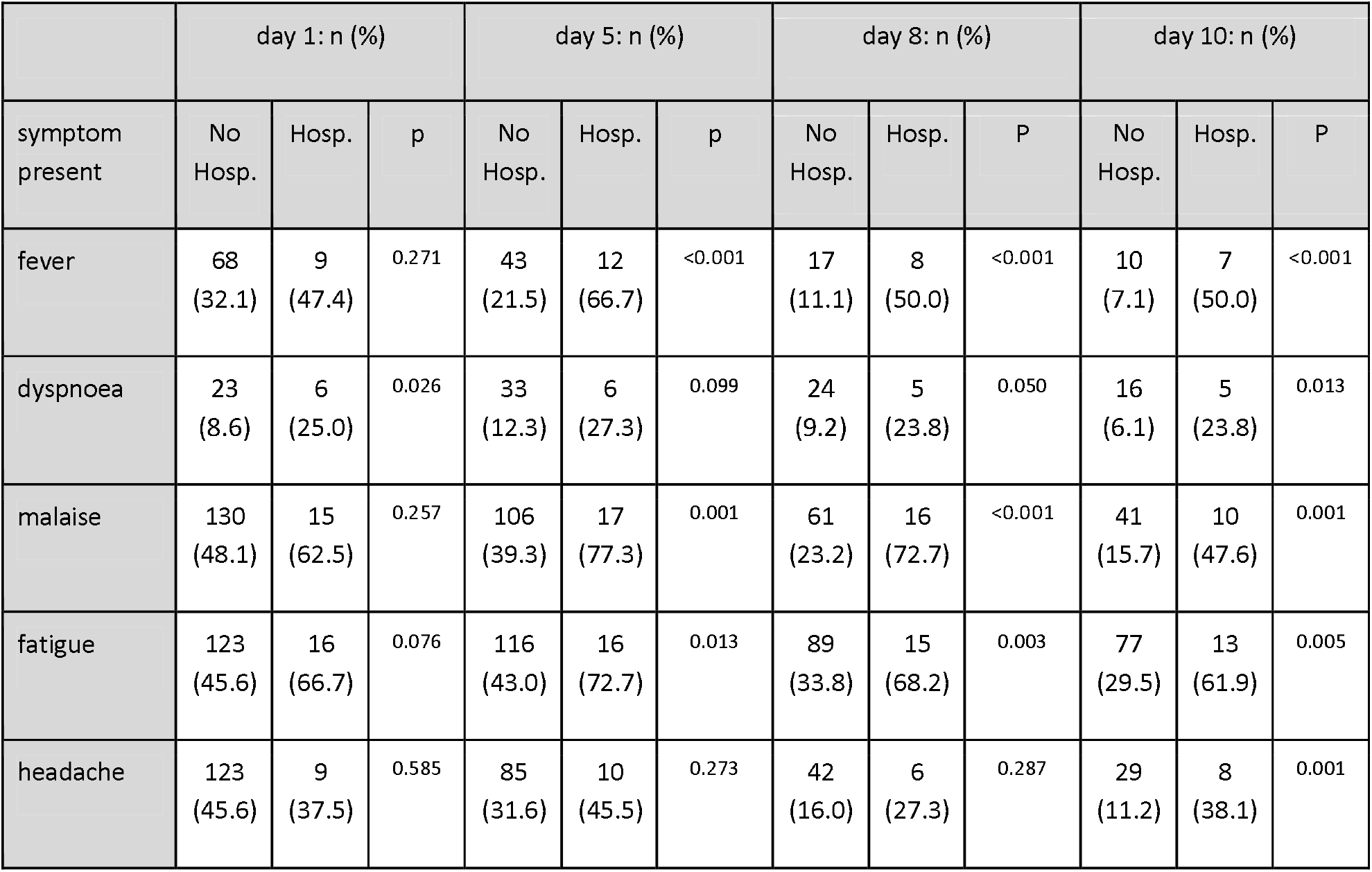
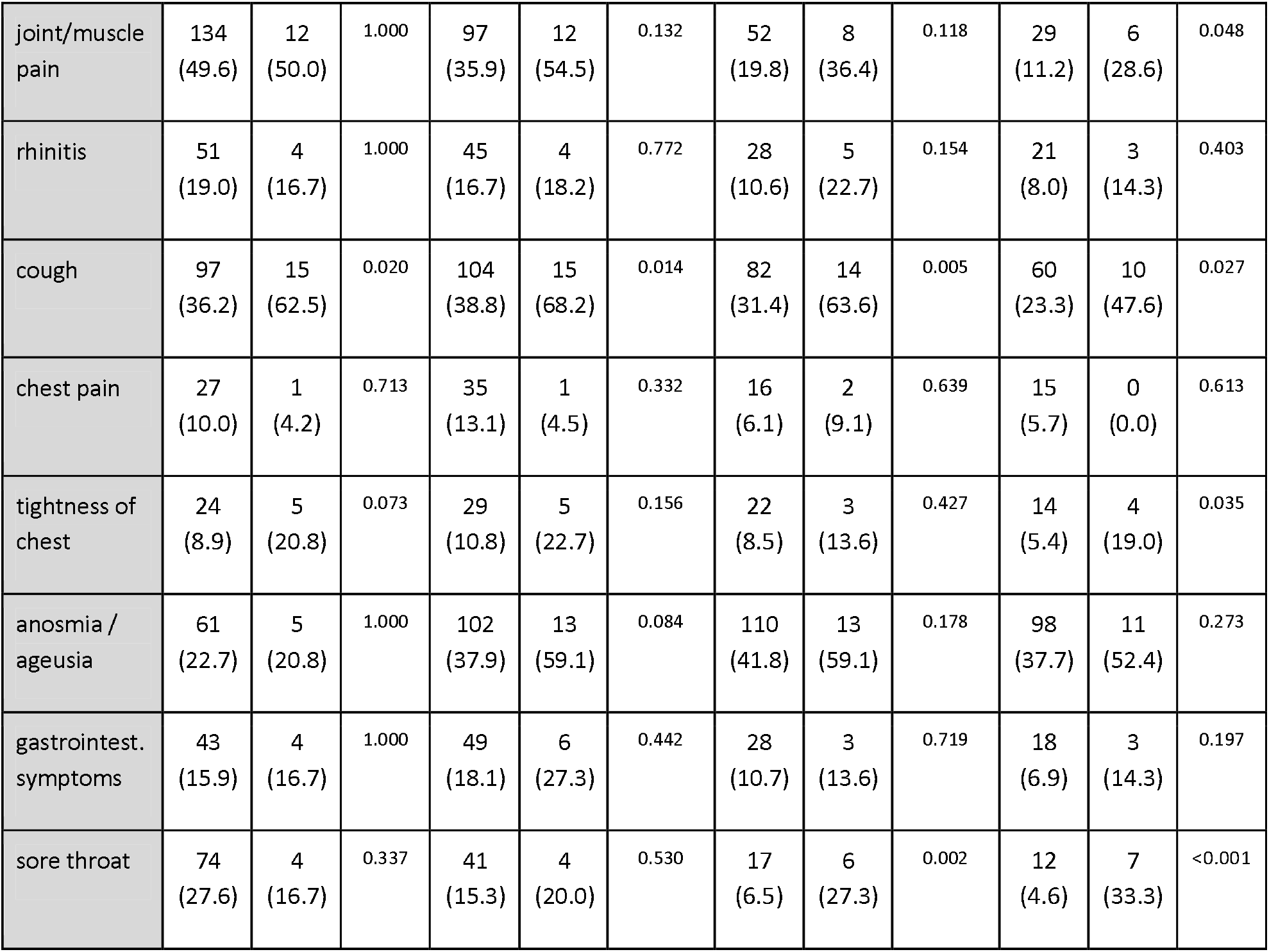
Hospital admission vs. non-admission in relation to the presence of a symptom

|  | day 1: n (%) |  |  | day 5: n (%) |  |  | day 8: n (%) |  |  | day 10: n (%) |  |  |
| --- | --- | --- | --- | --- | --- | --- | --- | --- | --- | --- | --- | --- |
| symptom present | No Hosp. | Hosp. | p | No Hosp. | Hosp. | p | No Hosp. | Hosp. | P | No Hosp. | Hosp. | P |
| fever | 68<br>(32.1) | 9<br>(47.4) | 0.271 | 43<br>(21.5) | 12<br>(66.7) | <0.001 | 17<br>(11.1) | 8<br>(50.0) | <0.001 | 10<br>(7.1) | 7<br>(50.0) | <0.001 |
| dyspnoea | 23<br>(8.6) | 6<br>(25.0) | 0.026 | 33<br>(12.3) | 6<br>(27.3) | 0.099 | 24<br>(9.2) | 5<br>(23.8) | 0.050 | 16<br>(6.1) | 5<br>(23.8) | 0.013 |
| malaise | 130<br>(48.1) | 15<br>(62.5) | 0.257 | 106<br>(39.3) | 17<br>(77.3) | 0.001 | 61<br>(23.2) | 16<br>(72.7) | <0.001 | 41<br>(15.7) | 10<br>(47.6) | 0.001 |
| fatigue | 123<br>(45.6) | 16<br>(66.7) | 0.076 | 116<br>(43.0) | 16<br>(72.7) | 0.013 | 89<br>(33.8) | 15<br>(68.2) | 0.003 | 77<br>(29.5) | 13<br>(61.9) | 0.005 |
| headache | 123<br>(45.6) | 9<br>(37.5) | 0.585 | 85<br>(31.6) | 10<br>(45.5) | 0.273 | 42<br>(16.0) | 6<br>(27.3) | 0.287 | 29<br>(11.2) | 8<br>(38.1) | 0.001 |
| joint/muscle pain | 134<br>(49.6) | 12<br>(50.0) | 1.000 | 97<br>(35.9) | 12<br>(54.5) | 0.132 | 52<br>(19.8) | 8<br>(36.4) | 0.118 | 29<br>(11.2) | 6<br>(28.6) | 0.048 |
| rhinitis | 51<br>(19.0) | 4<br>(16.7) | 1.000 | 45<br>(16.7) | 4<br>(18.2) | 0.772 | 28<br>(10.6) | 5<br>(22.7) | 0.154 | 21<br>(8.0) | 3<br>(14.3) | 0.403 |
| cough | 97<br>(36.2) | 15<br>(62.5) | 0.020 | 104<br>(38.8) | 15<br>(68.2) | 0.014 | 82<br>(31.4) | 14<br>(63.6) | 0.005 | 60<br>(23.3) | 10<br>(47.6) | 0.027 |
| chest pain | 27<br>(10.0) | 1<br>(4.2) | 0.713 | 35<br>(13.1) | 1<br>(4.5) | 0.332 | 16<br>(6.1) | 2<br>(9.1) | 0.639 | 15<br>(5.7) | 0<br>(0.0) | 0.613 |
| tightness of chest | 24<br>(8.9) | 5<br>(20.8) | 0.073 | 29<br>(10.8) | 5<br>(22.7) | 0.156 | 22<br>(8.5) | 3<br>(13.6) | 0.427 | 14<br>(5.4) | 4<br>(19.0) | 0.035 |
| anosmia / ageusia | 61<br>(22.7) | 5<br>(20.8) | 1.000 | 102<br>(37.9) | 13<br>(59.1) | 0.084 | 110<br>(41.8) | 13<br>(59.1) | 0.178 | 98<br>(37.7) | 11<br>(52.4) | 0.273 |
| gastrointest. symptoms | 43<br>(15.9) | 4<br>(16.7) | 1.000 | 49<br>(18.1) | 6<br>(27.3) | 0.442 | 28<br>(10.7) | 3<br>(13.6) | 0.719 | 18<br>(6.9) | 3<br>(14.3) | 0.197 |
| sore throat | 74<br>(27.6) | 4<br>(16.7) | 0.337 | 41<br>(15.3) | 4<br>(20.0) | 0.530 | 17<br>(6.5) | 6<br>(27.3) | 0.002 | 12<br>(4.6) | 7<br>(33.3) | <0.001 |

COVID-19 seems to start with several symptoms simultaneously: Nearly half of the participants reported 3-5 symptoms on day 1. 10 participants had not reported symptoms on day one, with 3 of them having become symptomatic by day 5, and 7 remaining asymptomatic (Table 4).

**Table 4:**
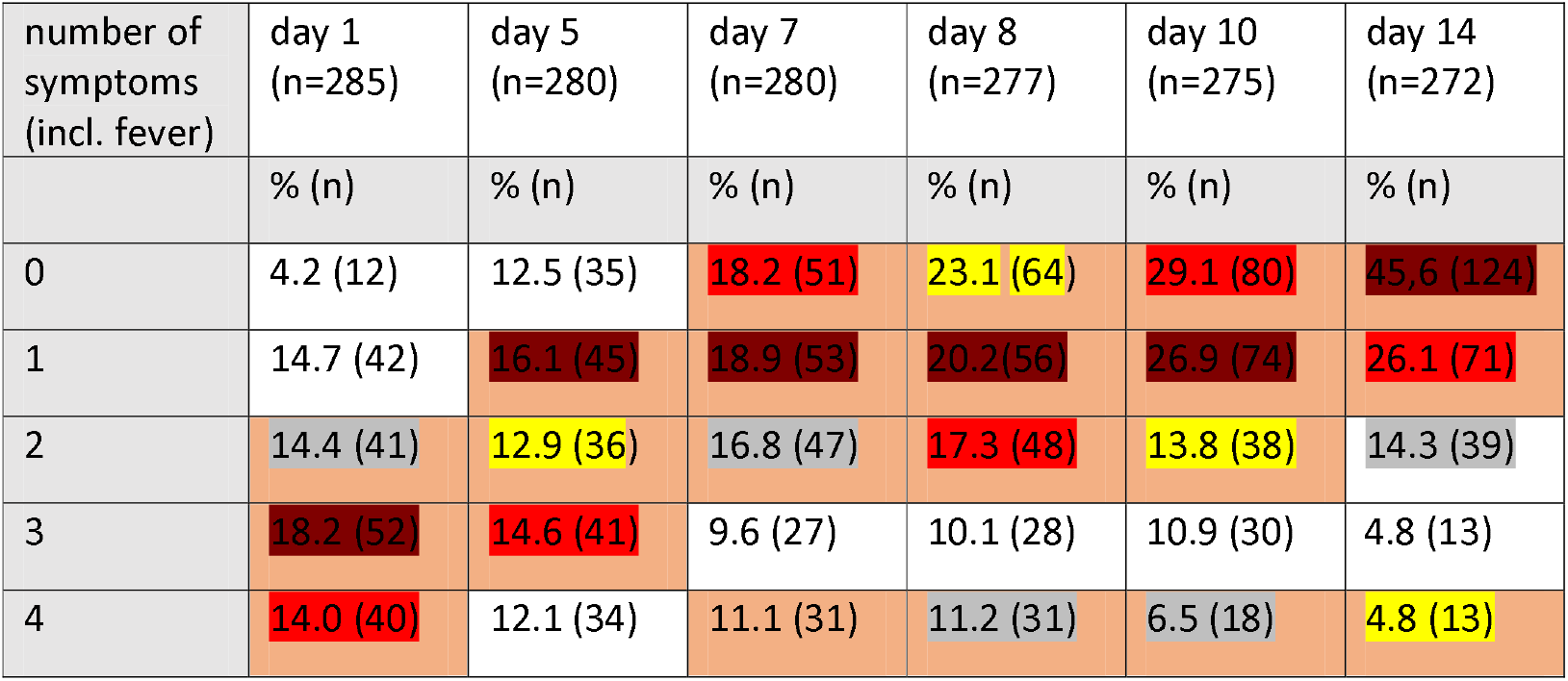
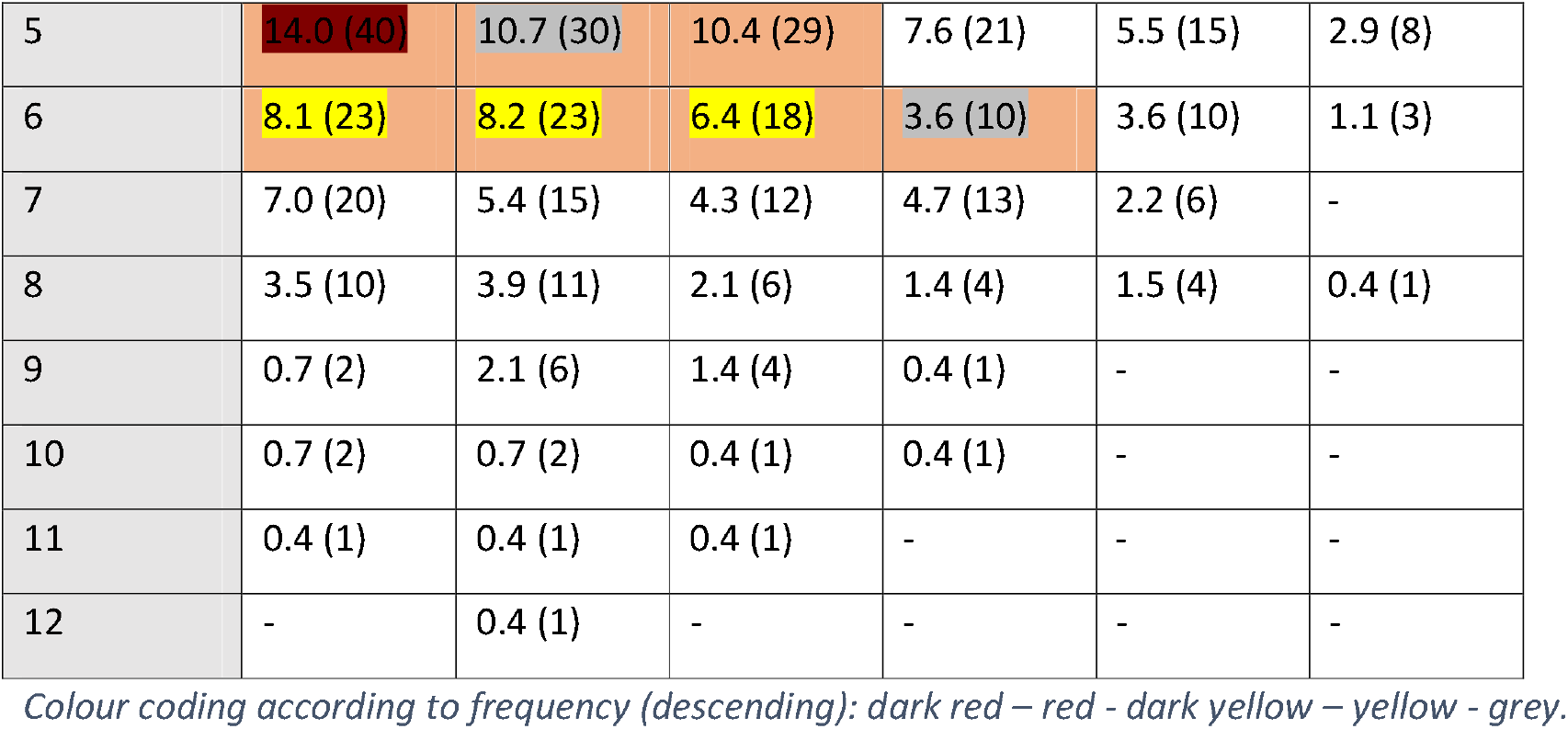
Number of symptoms by days 1,5,7,8,10,14.

| number of symptoms (incl. fever) | day 1 (n=285) | day 5 (n=280) | day 7 (n=280) | day 8 (n=277) | day 10 (n=275) | day 14 (n=272) |
| --- | --- | --- | --- | --- | --- | --- |
|  | % (n) | % (n) | % (n) | % (n) | % (n) | % (n) |
| 0 | 4.2 (12) | 12.5 (35) | 18.2 (51) | 23.1 (64) | 29.1 (80) | 45.6 (124) |
| 1 | 14.7 (42) | 16.1 (45) | 18.9 (53) | 20.2 (56) | 26.9 (74) | 26.1 (71) |
| 2 | 14.4 (41) | 12.9 (36) | 16.8 (47) | 17.3 (48) | 13.8 (38) | 14.3 (39) |
| 3 | 18.2 (52) | 14.6 (41) | 9.6 (27) | 10.1 (28) | 10.9 (30) | 4.8 (13) |
| 4 | 14.0 (40) | 12.1 (34) | 11.1 (31) | 11.2 (31) | 6.5 (18) | 4.8 (13) |
| 5 | 14.0 (40) | 10.7 (30) | 10.4 (29) | 7.6 (21) | 5.5 (15) | 2.9 (8) |
| 6 | 8.1 (23) | 8.2 (23) | 6.4 (18) | 3.6 (10) | 3.6 (10) | 1.1 (3) |
| 7 | 7.0 (20) | 5.4 (15) | 4.3 (12) | 4.7 (13) | 2.2 (6) | - |
| 8 | 3.5 (10) | 3.9 (11) | 2.1 (6) | 1.4 (4) | 1.5 (4) | 0.4 (1) |
| 9 | 0.7 (2) | 2.1 (6) | 1.4 (4) | 0.4 (1) | - | - |
| 10 | 0.7 (2) | 0.7 (2) | 0.4 (1) | 0.4 (1) | - | - |
| 11 | 0.4 (1) | 0.4 (1) | 0.4 (1) | - | - | - |
| 12 | - | 0.4 (1) | - | - | - | - |

Colour coding according to frequency (descending): dark red – red - dark yellow – yellow - grey.

68% of the patients reported symptoms persisting on day 10, and half the sample still had complaints on day 14.

A higher number of symptoms was associated with higher probability of hospital admission. This was significant for days 7 and 8 (Fig.2).

## Discussion

### Initial symptoms, and development of symptomatology

We found early symptoms to be mostly unspecific and often discreet, with joint or muscle pain, malaise and fatigue being the most common symptoms (Table 2, Fig.1). We did not find any of the 13 symptoms in the selection suitable to rule out or confirm early diagnosis of COVID-19 by clinical means alone, which is supported by several of the more recent studies based at least partly on data from primary care (7, 9).

Symptoms generally considered to increase likelihood for Covid diagnosis i.e. fever, dyspnoea and loss of taste and/or smell (7, 9, 10, 17, 18) we found to be comparatively rare, at least at early stages of the disease (Table 2, Fig.1). On day 1 less than 30% of patients reported fever, less than one third (25.9%) reported loss of smell or taste, and only 13.6 % suffered from dyspnoea which renders those symptoms not well suited as testing criteria. These results differ from other studies that had derived data from hospitalized patients exclusively, or in combination with self-reported data using symptom apps. All of them were conducted in patients that had been tested according to pre-established testing criteria (8, 10). Our findings suggest that neither presence nor absence of any symptom is suitable to rule out COVID-19, and confirm results from another study, showing that patients reporting any kind of symptoms of a broad range of diseases profit from testing without delay and without restrictions(19).

We found fever, cough and dyspnoea still considered relevant for case definitions and used as screening criteria in many countries. According to our findings this procedure could be an obstacle to correct early diagnosis and case finding. In our sample, none of those symptoms was experienced by more than slightly over a third of the patients at any time during the illness.

This finding may have some impact on testing strategies. Austria like other countries propagates low threshold testing by self testing kits, which mostly lack external validation of their sensitivity and specificity(20). Sensitivity varies considerably altogether in Lateral Flow Tests (LFT) (21). We found presence of any of the discreet and unspecific symptoms we investigated to possibly indicate Covid-19 infection and thus to increase pre-test probability - particularly as long as the prevalence of SARS-CoV-2 is clearly higher than that of any other respiratory virus. Especially self-administered tests may yield false negative results. Even antigen tests of higher sensitivity should be repeated if one of those symptoms is present since virus load can be too low for detection early into the disease. Users and decision makers need to be aware of this (22).

Anosmia is known to be the most specific symptom(8), but according to our results tends to appear in the later course of the disease. As an initial symptom we could trace it in only 22% of patients, but in double as many on days 7 and 8 (Table 2). Menni (8) identified this symptom in 64.5% of patients, and concludes that this symptom could help early diagnosis. Our findings do not support this conclusion. The difference may be caused by the time of detection or selection bias and thus shows the relevance of early investigation of symptoms at a low threshold point of care (13).

This finding should lead to reconsider contact-tracing strategies: finding the symptom anosmia might indicate delayed diagnosis, and should prompt an extension of the contact-tracing period to at least 7 days before the infection was detected.

### Associations between symptoms and hospitalization

We found several associations between symptoms and the need for hospitalization. Our findings suggest that dyspnoea is prone to lead to admission to hospital when present on day 1, as well as on day 10 and significantly on day 14. Fatigue as well as malaise through the whole course of the disease starting from day 5, and cough, persisting on day 8 seem to be associated with higher rates of hospitalization, fever only if present from day 5 on (Table 3, Fig.2). Dyspnoea has been shown to be predictive for hospital admission in one systematic review and a meta-analysis (2, 17), which is in line with our findings. No other predictive symptoms have consistently been identified so far. Our findings on persisting symptoms at the end of the observational period fit in well with recent studies on Long COVID-19 (23, 24)

We observed a higher number of symptoms (3 or more) on day 7 and 8 to be associated with a significantly higher probability of hospital admissions (Table 5). Patients in the non-hospitalized group experience their peak in number of symptoms on day 1 to day 5. Patients in the hospitalized group had more symptoms from the start and experienced a further rise on day 5 as well as a markedly slower decrease. This could indicate that patients might profit from being closely monitored: Finding a rise in number of symptoms should arouse suspicion of an imminent severe course of disease. This might, if corroborated, even allow identifying individual cut off points to introduce innovative early interventions presently under discussion like budesonide, low-molecular heparins or monoclonal antibody therapy (25, 26).

Over 50% of patients reported symptoms at the end of the observational period on day 14, and more than two thirds on day 10, when the isolation period usually ends. Mostly this concerns loss of taste or smell or fatigue. This coincides with the most common complaints reported by patients suffering from Long Covid (23). Regarding the fact that Long Covid is in many cases not a trivial complaint but may lead to delayed and severe complications (27), it seems justifiable to recommend medical examination after isolation to decide if and when a patient can be considered healthy and safe to return to physical activity and/or work.

### Strengths and limitations

This study to our knowledge is the first one to investigate data on the course of COVID-19 collected exclusively from patients in primary care. GPs were free to make their own testing decision according to their clinical judgement, and they followed each patient individually from day 1 to day 10 or 14 in most cases. Only 10% of the patients included in our study approached primary care after having been tested according to formal testing criteria by another testing facility. Most other studies (2, 7, 9, 10) recruited patients mainly or exclusively via symptom apps or in hospital care, after the testing decision had been made according to established testing criteria which makes them less likely to detect early symptoms not already known to be associated with Covid-19.

The study has several limitations though. We could recruit 25 practices in 7 out of the 9 Austrian provinces. The average number of patients included per practice was 12 (SD 8.9). The limited number of patients is probably due to the increased workload under difficult working conditions during the pandemic in combination with a rather extensive questionnaire and the need to follow patients over a longer period as well as the effort not being remunerated. We have to reckon with possible recruiting bias. However, the number of cases needed to identify group differences was calculated in advance, and this number has been reached. Our overall results are in accordance with our preliminary result analyses.

Another limitation is that not all data on symptoms were provided by the GPs, particularly on temperature. Most likely this applies mainly for symptoms which were not present, but this will have to be clarified by further research. Some patients (approximately 10%) were not diagnosed in primary care, so a possible confounder “testing criteria” cannot entirely be ruled out. We are likely to have overestimated some symptoms on day 1, because a proportion of patients may have unknowingly been diagnosed later than that.

We could not detect specific patterns of symptom-combinations. This may be due to the limited sample size.

## Conclusion

We could demonstrate a variety of unspecific symptoms to be clearly more common - particularly in the early stages - than those widely considered typical of Covid-19. Understanding this can help to avoid missing potentially infectious patients because they are either not tested at all, or because inappropriate testing methods like low sensitivity self-test kits are being used. Low threshold contact for patients with any kind of symptoms combined with testing at the point of care can accelerate diagnosis and enhance effective contact tracing.

We found several symptoms possibly indicating future complications. This knowledge in conjunction with timely identification in primary care could help averting severe disease in some cases. To facilitate follow up in primary care, patients need to be either diagnosed there, or to reliably report to their GP if tested positive.

A high percentage of patients with Covid-19 experience symptoms for more than 10 or even 14 days. A final check-up should be recommended at least for those who do not feel fully fit before they plan to return to physical and occupational activities.

### Implications for further research

Data collected in primary care settings can provide additional information and can offer a wider spectrum of understanding COVID-19 disease. The results of our exploratory retrospective study should be controlled by a prospective study.

We did not record data on patients who tested negative or were not tested at all. Future investigations into this topic should aim at recording symptoms in all patients reporting for suspected SARS-CoV-2 infection.

## Data Availability

All data referred to in the manuscript are available from:
Department of General Medicine and Family Practice, Karl Landsteiner Privatuniversitaet, Krems, Austria

## Additional information

### Ethical approval

was given by Commission for Scientific Integrity and Ethics, Karl-Landsteiner Private University for Health Sciences, Krems, Austria. EK Nr: 1046/2020 13/07/20

The study was registered as DRKS00022448 on DRKS

## Acknowledgements

The Austrian Society of General Practice and Family Medicine provided support by assisting the process of recruitment of study practices. This was essential for this study to be undertaken.

We thank all our colleague GPs who undertook the task of recruiting patients, completing questionnaires and onward them to us.

And we thank the many patients, who contributed their data and recorded their symptoms.

---

**Table S1:**
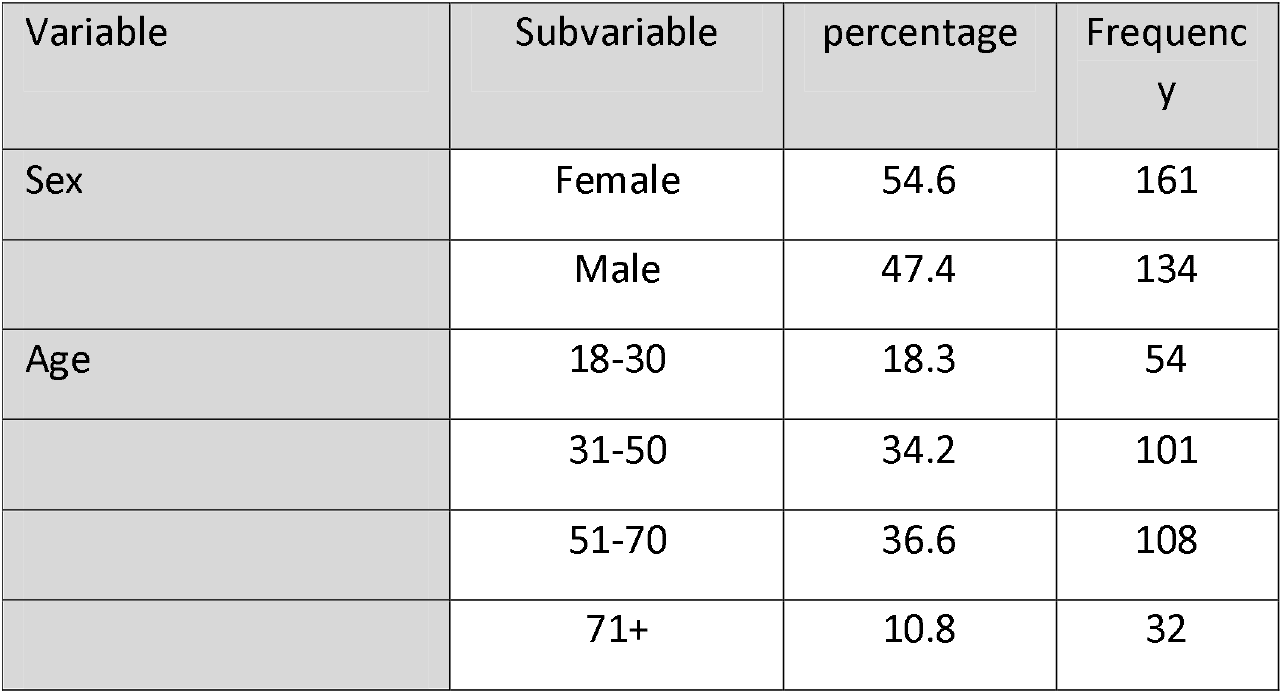
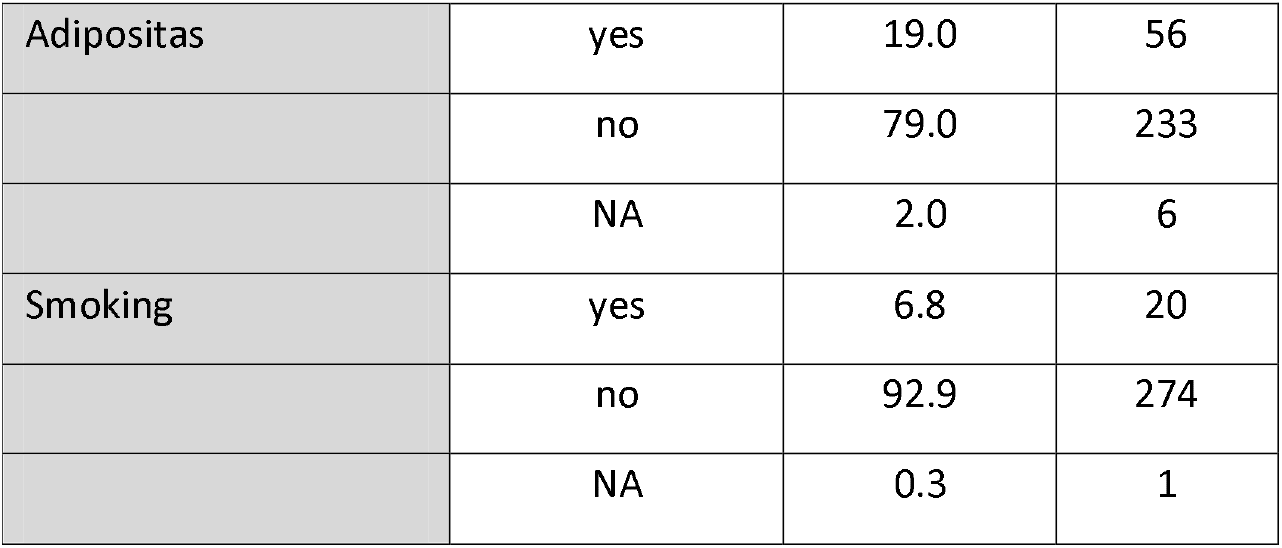
Demographics (N=295)

| Variable | Subvariable | percentage | Frequenc<br>y |
| --- | --- | --- | --- |
| Sex | Female | 54.6 | 161 |
|  | Male | 47.4 | 134 |
| Age | 18-30 | 18.3 | 54 |
|  | 31-50 | 34.2 | 101 |
|  | 51-70 | 36.6 | 108 |
|  | 71+ | 10.8 | 32 |
| Adipositas | yes | 19.0 | 56 |
|  | no | 79.0 | 233 |
|  | NA | 2.0 | 6 |
| Smoking | yes | 6.8 | 20 |
|  | no | 92.9 | 274 |
|  | NA | 0.3 | 1 |

**Table S2:**
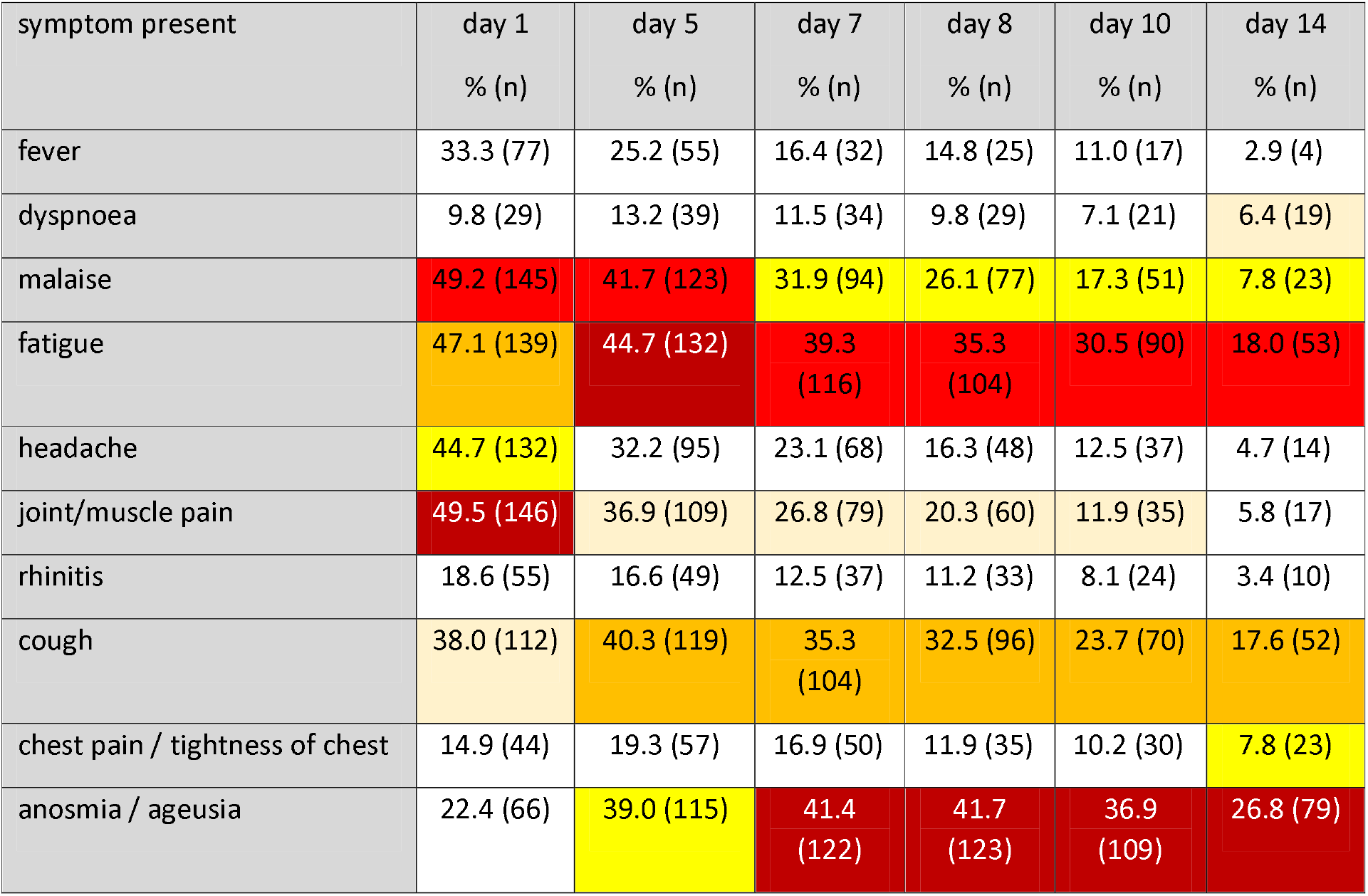
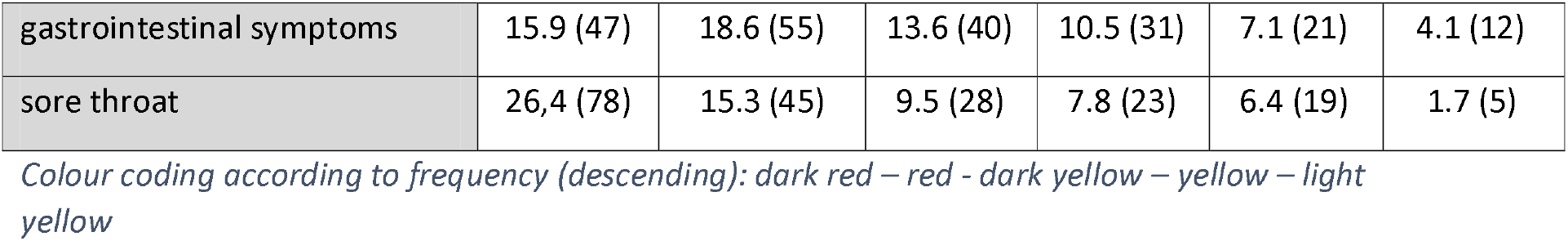
Symptoms reported on day 1,5,7,8,10,14 (N=295)

| symptom present | day 1 | day 5 | day 7 | day 8 | day 10 | day 14 |
| --- | --- | --- | --- | --- | --- | --- |
|  | % (n) | % (n) | % (n) | % (n) | % (n) | % (n) |
| fever | 33.3 (77) | 25.2 (55) | 16.4 (32) | 14.8 (25) | 11.0 (17) | 2.9 (4) |
| dyspnoea | 9.8 (29) | 13.2 (39) | 11.5 (34) | 9.8 (29) | 7.1 (21) | 6.4 (19) |
| malaise | 49.2 (145) | 41.7 (123) | 31.9 (94) | 26.1 (77) | 17.3 (51) | 7.8 (23) |
| fatigue | 47.1 (139) | 44.7 (132) | 39.3 (116) | 35.3 (104) | 30.5 (90) | 18.0 (53) |
| headache | 44.7 (132) | 32.2 (95) | 23.1 (68) | 16.3 (48) | 12.5 (37) | 4.7 (14) |
| joint/muscle pain | 49.5 (146) | 36.9 (109) | 26.8 (79) | 20.3 (60) | 11.9 (35) | 5.8 (17) |
| rhinitis | 18.6 (55) | 16.6 (49) | 12.5 (37) | 11.2 (33) | 8.1 (24) | 3.4 (10) |
| cough | 38.0 (112) | 40.3 (119) | 35.3 (104) | 32.5 (96) | 23.7 (70) | 17.6 (52) |
| chest pain / tightness of chest | 14.9 (44) | 19.3 (57) | 16.9 (50) | 11.9 (35) | 10.2 (30) | 7.8 (23) |
| anosmia / ageusia | 22.4 (66) | 39.0 (115) | 41.4 (122) | 41.7 (123) | 36.9 (109) | 26.8 (79) |
| gastrointestinal symptoms | 15.9 (47) | 18.6 (55) | 13.6 (40) | 10.5 (31) | 7.1 (21) | 4.1 (12) |
| sore throat | 26,4 (78) | 15.3 (45) | 9.5 (28) | 7.8 (23) | 6.4 (19) | 1.7 (5) |

Colour coding according to frequency (descending): dark red – red - dark yellow – yellow – light yellow

**Figure S1.**
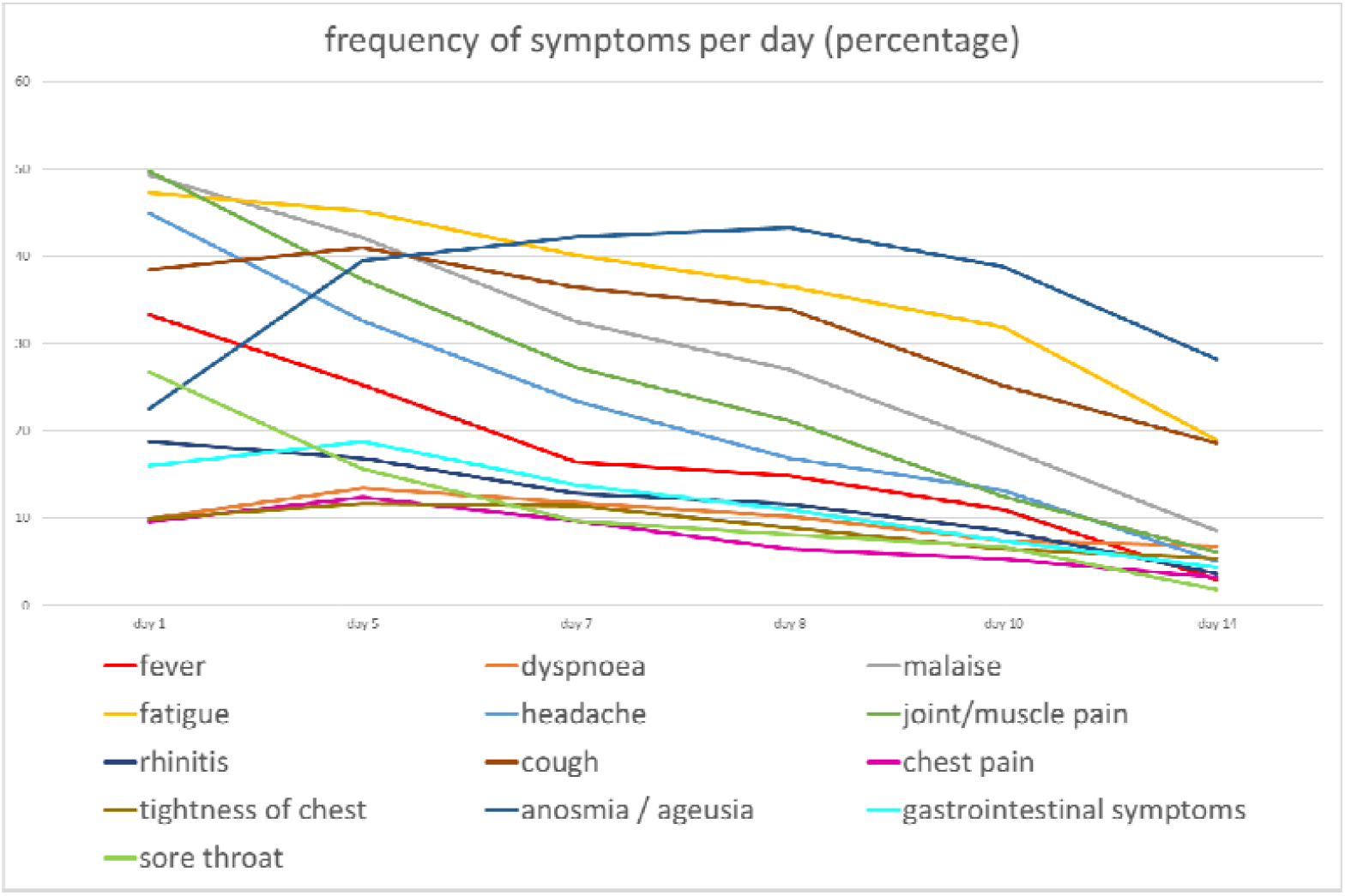
Frequencies of symptoms in the course of the disease

